# Factors influencing COVID rates at local authority level and contribution of variation in vaccine coverage

**DOI:** 10.1101/2021.03.08.21253144

**Authors:** Rachel Cullum, Padmanabhan Badrinath

## Abstract

We have undertaken a piece of rapid analysis of the most recent COVID-19 weekly rates to examine commonalities across areas with rates exceeding twice the national average. Our preliminary findings point towards an association between higher case rates and deprivation with implications for health inequalities. Furthermore, we also observed an association between higher case rates and lower rates of vaccination. More analysis is needed to further explore these linkages and help fuel what should be an urgent narrative around the need to tackle health inequalities in the COVID era.

## Introduction

It is promising and positive news that funding is being provided to local areas to reduce vaccine inequalities (1). This is a welcome step for attempting to reduce some of the health inequalities associated with COVID-19. However, vaccination rates are not the only variable associated with the variation in COVID-19 rates across local authority areas in England.

We have undertaken some rapid analysis of the latest weekly COVID-19 rates (4 March 2021) at lower tier authority level (2). Our aim was to explore some of the commonalities across areas that have more than twice the average overall population COVID19 rate in England or twice the rate for over 65s which were 103 cases per 100,000 and 69 cases per 100,000 respectively.

## Findings

We have used the latest population data (mid-2019) (3) to estimate the proportion of the population currently vaccinated with at least one dose across lower tier authority areas. We must insert a caveat here – these percentages are based on mid-2019 data, which will clearly be out of date. However, if we assume steady population growth, it is still valuable to use these rates for comparative purposes.

It appears that there are differences in vaccination rates between areas with higher and lower numbers of cases (4). On average, in areas with COVID-19 rates ≥206 per 100,000, the population vaccination rate is 30.3%. Across all other lower tier authorities, the average is 31.85%. We also looked at the corresponding statistics for over-65s. In areas with COVID-19 rates of ≥139 per 100,000 in over-60s (over twice the case rate for England for over-60s), on average 92.27% of the population aged over 65 has been vaccinated. Across all other areas, the average is 93.46%.

However, when it comes to comparative deprivation, the results are starker.

In order to compare COVID-19 rates with deprivation rates in lower tier areas, we used the most recent (2019) Index of Multiple Deprivation (IMD) data, which contains a subset looking of domains relating to health deprivation and disability (5). In areas with COVID-19 rates ≥206 per 100,000, the average IMD score is 24, while in the remaining areas* the average score is 20 (4). The difference is more remarkable when it comes to those areas with higher rates of COVID-19 in the over-60s. In areas with COVID-19 rates of ≥139 per 100,000 in over-60s, the average IMD score is 28, while the average for all other areas is 19.

The same disparity can be seen when we only look at those IMD domains that relate to health deprivation and disability. The scale for this runs from – 2.63 in the Isles of Scilly (the best scoring area) up to 1.64 in Blackpool (the worst scoring area) (5). Across areas with rates ≥206 per 100,000, the average score is 0.35. Across all other areas, the average score is −0.14. Again, the difference is more noticeable when we examine the disparity between areas with higher and lower rates in the over-60s age group. In areas with COVID-19 rates of ≥139 per 100,000 in over, the average IMD score for health, deprivation and disability is 0.45. Across all other areas, the average is −0.15.

We examined test rates as a potential alternative explanation for differences in COVID-19 rates across areas and found that there are differences between the average number of tests in higher and lower case rate areas (6). In areas with COVID-19 rates ≥206 per 100,000, the average number of tests per 100,000 population is 396; in the remainder of areas, it is 369.

However, this pattern is not reflected when we look at areas with higher case rates in the over 60s. In those areas, the average number of individuals tested per 100,000 people is 363, vs 369 across the other areas. Additionally, none of the areas we looked at (the highest case rate areas for the whole population or the over 60s) fell into the list of top 25 areas for testing rate per 100,000 people.

Furthermore, there seems to be something of a north-south divide. Out of the eight areas with rates ≥206 per 100,000, 75% of these are in the Midlands (4 in the East Midlands, 2 in the West Midlands) with the remaining two areas situated in the East of England. None are in London, which is interesting and unexpected given that London is such a large urban centre with complex transport systems and plenty of potential for transmission. Of the 11 areas with rates of ≥139 per 100,000 in over-60s, 63% (7) are in the Midlands and 27% (3) are in the North (North West and Yorkshire). Again, none are in London, although one area is in the South East (Slough). The latest data available show that between 22^nd^ – 28^th^ February 2021, COVID-19 admissions to hospital and diagnoses in hospital were highest in the Midlands (1,368), the North East and Yorkshire (1,126) and the North West (857) (7).

## Discussion

Our findings carry a worrying message around health inequalities. The BBC has recently highlighted that there is a risk that COVID-19 will become a ‘disease of the poor’ (8). Their report highlights that people in poorer areas are less likely to have the vaccine, and that there is a higher death rate from COVID-19 in poorer areas of England. In this report, the Director of Public Health for Lancashire stated that the combination of lower vaccine uptake and higher infection levels in poorer areas could result in inequalities perpetuating for years to come. Furthermore, the recently published Marmot COVID Review (9) focused on COVID-19 concluded that there is a high mortality rate in Black, Asian, and ethnic minority groups because of structural racism in society leading to adverse social and economic conditions for these groups.

When considering disparity in COVID-19 rates across areas, we must not fail to recognise that there are structural societal inequalities operating here. Work is needed to tackle these deep-seated health inequalities that perpetuate with each intergenerational cycle. We note with interest that the Association of Directors of Public Health (ADPH) has recently drawn attention to the need to address health inequalities as part of the new Health and Care Bill (10). As the UK exits the pandemic and move into the endemic phase and the public health system gets back to ‘business as usual’, a renewed focus on health inequalities would be a positive and energising starting point.

*Buckinghamshire has been removed from the analysis as there are no IMD data. Furthermore, some areas for which IMD data are provided have no corresponding COVID-19 data (Aylesbury Vale, Chiltern, Wycombe, and South Bucks).

## Data Availability

All the data for this research was obtained by publicly available information and some of the links are included below

https://www.gov.uk/government/publications/coronavirus-cases-by-local-authority-epidemiological-data-4-march-2021

https://www.england.nhs.uk/statistics/statistical-work-areas/covid-19-vaccinations/

https://opendatacommunities.org/data/societal-wellbeing/imd2019/indicesbyla

## Notes

### Competing Interest Statement

The authors work in a Local Authority Public Health Department. PB leads the local COVID specialist response team. The views are of the authors and in no way represent that of their employer Suffolk County Council

### Funding Statement

No funding was used for this research.

### Author Declarations

Ethical approval not needed as this research is based on publicly available data

